# Chronic Maternal White Blood Cell Elevation During Pregnancy and Offspring Autism

**DOI:** 10.64898/2026.11.15.26368136

**Authors:** Alon Tzoref, Amit Dichter, Gal Meiri, Idan Menashe

## Abstract

**Background:** Maternal immune activation is a robust risk factor for autism spectrum disorder (ASD). Current evidence relies on specialized immunological markers not routinely measured, limiting population-level risk identification. We investigated the association between maternal white blood cell (WBC) counts, a ubiquitous component of routine prenatal care during pregnancy, and offspring ASD risk.

**Methods:** We conducted a retrospective population-based cohort study of 303,234 singleton live births (51% male) between 2011 and 2021 from Clalit Health Services in Israel. Exposures were maternal total and differential WBC counts from routine testing during the 1st and 2nd trimesters. High and low exposures were defined as >97.5^th^ and <2.5^th^ percentiles, respectively. The primary outcome was clinical ASD diagnosis. Analyses included multivariable logistic regressions, matched-sibling analyses (n=2,894 discordant pairs), and chronicity analyses in multiparous mothers (n=72,102).

**Results:** Among 303,234 children, 8,106 (2.7%) were diagnosed with ASD. Elevated maternal total WBC counts were associated with increased odds of ASD (adjusted odds ratio [aOR]=1.59; 95% CI, 1.43-1.75), with the strongest association in the 1st trimester (aOR=1.64; 95% CI, 1.45-1.85). Similar associations were seen with elevated neutrophils, lymphocytes, and monocytes. While most associations were attenuated in matched-sibling analyses, chronicity analysis revealed increased ASD odds in offspring of mothers with persistently elevated WBC counts across multiple pregnancies (aOR=1.49; 95% CI, 1.08-2.01).

**Conclusions:** Persistently elevated maternal WBC counts across pregnancies are associated with increased offspring ASD risk. These routinely collected hematologic markers support the maternal immune activation hypothesis and may contribute to future prenatal risk assessment models.

## Introduction

Autism spectrum disorder (ASD) is a complex neurodevelopmental condition with a rising global prevalence and a multifactorial etiology involving genetic and environmental factors.(1) Accumulating evidence highlights the prenatal period, particularly early gestation, as a critical window of vulnerability where various intrauterine exposures can disrupt fetal brain development.(2-4) Identifying such prenatal risk factors for ASD may facilitate early risk stratification, ensuring that high-risk infants are identified promptly to benefit from effective early-life interventions. (5-9)

Maternal immune dysregulation during gestation has emerged as a risk factor contributing to altered fetal neurodevelopment and consequently the diagnosis of ASD in the offspring.(10-12) Previous studies and meta-analyses have linked maternal autoimmune diseases, prenatal infections, and elevated inflammatory biomarkers such as C-reactive protein (CRP) to an increased risk of ASD in the offspring .(13-15) Mechanistic studies further suggest that this effect is mediated by pro-inflammatory cytokines such as Interleukin-6 (IL-6) and Interleukin-17A (IL-17A), which can cross the placenta and alter brain development. (16-19) Nevertheless, the relatively high cost of cytokines and autoantibodies tests and the lack of standardized reference ranges for pregnant populations, limit their clinical utility for large-scale risk identification.

Alternatively, white blood cell (WBC) count is a ubiquitous, cost-effective component of routine prenatal screening that may serve as a reliable indicator for maternal inflammatory status. Yet, to the best of our knowledge, prior research in the context of offspring neurodevelopment has focused largely on secretory products or clinical inflammatory conditions rather than cellular counts.

Furthermore, no studies to date have systematically examined whether routine, asymptomatic variations in maternal WBC profiles during specific gestational windows are associated with offspring ASD risks. Moreover, it remains unclear whether these associations reflect transient acute responses or a stable maternal immune phenotype persisting across pregnancies.

To address these research gaps, we conducted a population-based cohort study of over 300,000 births from a single health maintenance organization (HMO) in Israel and tested the association between trimester-specific maternal WBC counts and ASD risk in offspring. Furthermore, we leveraged the significant portion of multiparous women in the cohort and matched-sibling analysis to examine whether extreme WBC counts are associated with ASD independent of shared familial and genetic confounders, and whether they reflect transient acute or a stable maternal immune condition.

## Methods and Materials

### Study Design and Population

This retrospective cohort study utilized data from Clalit Health Services (CHS), the largest healthcare provider in Israel, covering approximately 4.6 million members. The study population comprised all singleton live births > 25 weeks of gestation to Jewish mothers insured by CHS between January 1, 2011, and December 31, 2021. The analysis was restricted to the Jewish population to ensure socioeconomic and clinical homogeneity, as well as to mitigate substantial missing data in other sub-populations resulting from lower baseline utilization of routine prenatal care.(20) Exclusion criteria included: known genetic abnormalities associated with ASD (e.g., Fragile X, Down syndrome, Tuberous sclerosis), and absence of maternal blood count measurements during the 1^st^ or 2^nd^ trimester. To ensure data completeness and adequate outcome tracking, inclusion required a minimum of 3 years of follow-up from birth, which is clinically sufficient to capture the vast majority of ASD diagnoses in Israel.(21) After applying these criteria, the final study population consisted of 303,234 mother-child pairs, of which 8,106 children (2.7%) were diagnosed with ASD (detailed flowchart in **Figure 1)**.

**Figure 1:**
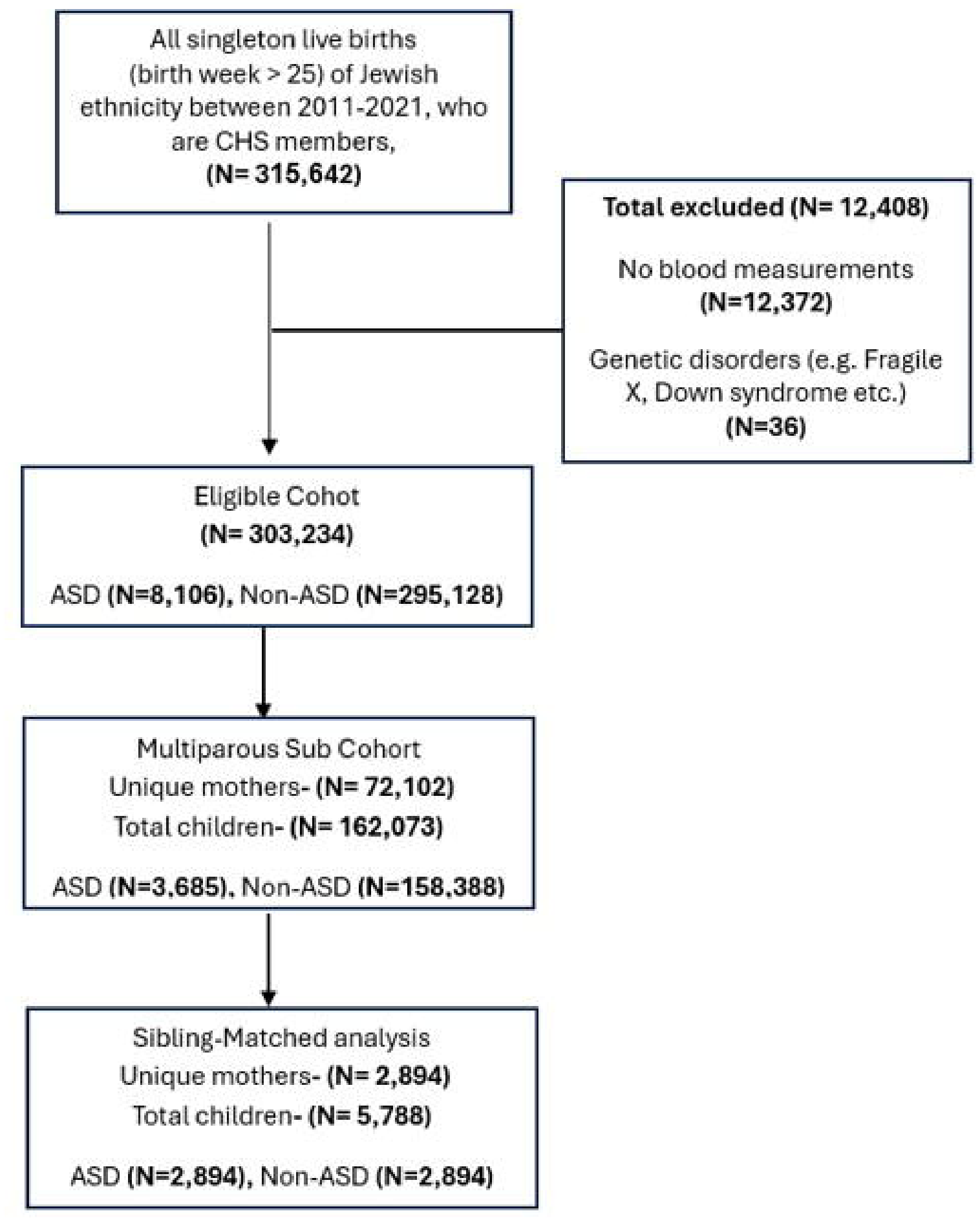
Flowchart of study cohort ascertainment.

### Data Collection

Data were extracted from the CHS electronic database via the MDClone system and analyzed within CHS’s secure Virtual Desktop Infrastructure (VDI) environment. The dataset incorporated a broad array of maternal and child characteristics, laboratory results, and clinical outcomes. To ensure data integrity, potential confounders with more than 20% missing values were excluded, and categorical variables with inconsistent labeling were harmonized by re-coding conflicting entries into standardized, clinically defined categories.

Maternal total and differential WBC counts were obtained from routine complete blood count (CBC) tests performed during the 1^st^ (weeks 0 to 13+6 days) and 2^nd^ (weeks 14 to 26+6 days) trimesters. Laboratory values were filtered according to physiologically plausible limits to mitigate recording errors, in line with established clinical literature.(22-25) Biologically plausible extreme values within these limits were retained, as they formed the categorical basis of the exposure definition.

### Exposure Assessment

The primary exposure variables were maternal total WBC counts and counts of specific WBC types (neutrophils, lymphocytes, monocytes, eosinophils, and basophils). Given the physiological shifts during pregnancy and the lack of universally established reference ranges, maternal WBC counts were categorized rather than modeled continuously to align with clinical decision-making processes, where diagnostic and therapeutic interventions are typically triggered by laboratory results that fall outside physiological reference ranges, regardless of the specific numerical value. For each cell type, “High” and “Low” levels were defined as the top and bottom 2.5% of the distribution, respectively, in either the 1^st^ or 2^nd^ trimesters, with the middle 95% serving as the “Normal” reference group. The absolute numerical values corresponding to these trimester-specific percentiles are detailed in **supplementary Table 1**.

**Table 1:** Sociodemographic, maternal, and delivery characteristics of the study groups-exposure during the entire pregnancy.

| **Variable** |  | **Maternal WBC levels** |  |  | **P-value a vs. b** | **P-value a vs. b** |
|---|---|---|---|---|---|---|
|  |  | **Normal<sup>a</sup> (ref)<br>(N= 282,209)** | **Low<sup>b</sup><br>(N=10,242)** | **High<sup>c</sup><br>(N= 10,783)** |  |  |
| Socio-demographic characteristics |  |  |  |  |  |  |
| **Maternal age<br>[mean (+ SD)]** |  | 31.6 (±5.1) | 31.8 (±5.1) | 31.7 (±5.3) | 0.006 | 0.13 |
| **Offspring sex<br>[N(%)]** | **Male** | 145,035 (51%) | 5,199 (51%) | 5,478 (51%) | 0.2 | 0.2 |
|  | **Female** | 137,174 (49%) | 5,043 (49%) | 5,305 (49%) |  |  |
| **Socioeconomic status<br>[N(%)]** | **Very low** | 3,415 (1.3%) | 126 (1.3%) | 64 (0.6%) | <0.001 | <0.001 |
|  | **Low** | 54,502 (21%) | 2,767 (28%) | 1,803 (18%) |  |  |
|  | **Medium** | 98,797 (37%) | 4,067 (42%) | 3,772 (37%) |  |  |
|  | **High** | 84,456 (32%) | 2,205 (23%) | 3,482 (34%) |  |  |
|  | **Very high** | 24,284 (9.1%) | 592 (6.1%) | 1,009 (10.0%) |  |  |
| Maternal characteristics |  |  |  |  |  |  |
| **Maternal BMI<br>[mean (+ SD)]** |  | 22.1 (±4.3) | 21.4 (±4.1) | 22.9 (±4.7) | <0.001 | <0.001 |
| **Gestational diabetes<br>[N(%)]** |  | 14,721 (5.2%) | 335 (3.3%) | 1,239 (11%) | <0.001 | <0.001 |
| **Gestational hypertension<br>[N(%)]** |  | 9,814 (3.5%) | 296 (2.9%) | 782 (7.3%) | 0.001 | <0.001 |
| **Gestational anemia<br>[N(%)]** |  | 42,449 (15%) | 1,987 (19%) | 1,624 (15%) | <0.001 | >0.9 |
| **IVF<br>[N(%)]** |  | 10,561 (3.7%) | 329 (3.2%) | 643 (6.0%) | 0.005 | <0.001 |
| Delivery characteristics |  |  |  |  |  |  |
| **Birth weight (gr)<br>[mean (+ SD)]** |  | 3,287 (±432) | 3,257 (±436) | 3,224 (±463) | <0.001 | <0.001 |
| **Birth week<br>[mean (+ SD)]** |  | 39.15 (±1.40) | 39.29 (±1.42) | 38.81 (±1.51) | <0.001 | <0.001 |

<sup>a</sup> WBC level ≥2.5% or ≤97.5% of the distribution; <sup>b</sup> WBC level <2.5% of the distribution; <sup>c</sup> WBC level >97.5% of the distribution

### Outcome Definition

The primary outcome was a diagnosis of ASD in the offspring, identified via CHS computerized medical records using ICD codes (ICD-9 code 299.0, autistic disorder). Because documentation of an ASD diagnosis according to DSM-5 criteria is a prerequisite for state-funded health services and benefits, the registry captures the vast majority of formal diagnoses. The study’s 3-year follow-up criterion ensures adequate time to capture these outcomes, as the median age of ASD diagnosis in Israel falls well within early childhood.(21)

### Covariates

Potential confounders included sociodemographic factors (maternal age, socioeconomic status, offspring sex), maternal characteristics (BMI, gestational diabetes, hypertension, anemia, IVF conception), and perinatal factors (birth weight, gestational age at delivery). All included variables met the pre-specified criterion of less than 20% missing data.

### Statistical Analysis

Descriptive statistics characterized the study population. Univariable analyses (Chi-square, t-tests) used to test the significance of background characteristic differences between exposure groups. The primary analysis employed multivariable logistic regression models to estimate adjusted odds ratios (aORs) and 95% confidence intervals (CIs) for the association between maternal WBC levels and ASD, adjusting for all potential confounders associated with both exposure and outcome variables. In the primary analysis, women with “low” or “high” WBC values in either the 1^st^ or 2^nd^ trimesters were aggregated into their respective exposure groups. Separate models were also constructed to test exposures restricted to the 1^st^or 2^nd^ trimesters.

To investigate the role of shared familial and genetic factors, a matched-sibling analysis was conducted comparing children with ASD to a randomly selected unaffected sibling using conditional logistic regression. This conditional model adjusted exclusively for pregnancy-specific, time-varying covariates, while time-invariant maternal characteristics were naturally conditioned out by the sibling fixed-effects design.

To determine whether the observed associations reflected a stable maternal immune condition across reproductive lifespans rather than transient acute responses, a chronicity analysis was performed to assess the risk of ASD in subsequent offspring. For that, a sub-cohort of all multiparous mothers who had at least two documented pregnancies within the registry (162,073 total pregnancies) was used. Mothers were considered to have a chronic high level if their WBC counts consistently fell within the top 97.5^th^ percentile across all of their documented pregnancies. We also used a cut-off point of 95^th^ percentile for this analysis increase the size of the exposure group. We compared women in the “chronically high” group to mothers whose counts were consistently within the normal range throughout all pregnancies. Here too, separate analyses were conducted for 1^st^ trimester, 2^nd^ trimester, and either 1^st^ or 2^nd^ trimester respectively.

All statistical analyses were performed using R statistics (version 4.2.2). A p-value <0.05 was considered statistically significant.

### Ethical Approval

The study protocol was approved by the Institutional Review Board of Soroka University Medical Center (SOR-237-21). The requirement for informed consent was waived due to the retrospective nature of the study and the use of anonymized health records.

## Results

The study eligible cohort comprised of 303,234 singleton live births of Jewish ethnicity, born after 25 weeks of gestation with available blood count data and no known genetic syndromes associated with ASD **(Figure 1)**. Of these, 162,073 births belong to 72,102 multiparous mothers during the study period. A total of 8,106 children in the total cohort (2.7%) and 3,685 children in the multiparous sub cohort (2.3%) were diagnosed with ASD.

Maternal WBC measurements were available for 256,548 pregnancies during the 1^st^ trimester and 235,490 during the 2^nd^ trimester. The distributions of maternal WBC counts are presented in **Figure S1 and Figure S2**. Total WBC, neutrophil, and lymphocyte counts followed an approximately normal distribution, while monocytes, eosinophils, and basophils exhibited right-skewed distributions, characterized by concentration near lower values and long tails toward higher values. Longitudinal analysis between the 1^st^ and 2^nd^ trimesters revealed moderate-to-strong intra-individual correlations across all cell types (Pearson’s r range: 0.34 [Basophils] - 0.68 [Eosinophils]; **Figure S3)**. This consistency indicates that maternal immune status remains relatively stable throughout pregnancy, justifying the assessment of both trimester-specific and combined exposure models.

Baseline sociodemographic and clinical characteristics of the study population, stratified by maternal WBC levels, are presented in **Table 1**. Although most group differences were statistically significant because of the large sample size, notable differences included the elevated gestational diabetes, hypertension, and IVF conception in the “high” WBC group and the lower socioeconomic status and higher rates of gestational anemia among women in the “low” WBC group. The same baseline characteristics were compared between ASD and non-ASD cases **(Table S2)**. As expected, the ASD group was characterized by a higher proportion of males (76% vs. 51%), advanced maternal age, and higher rates of maternal comorbidities and perinatal complications.

### Association Between Maternal WBC Counts and ASD

The results of the multivariable logistic regression analysis are summarized in **Table 2**. Elevated maternal total WBC counts (>97.5^th^ percentile) at any point during pregnancy were significantly associated with an increased risk of ASD in offspring (adjusted OR=1.59, 95% CI:1.43-1.75). When stratified by trimester, the association was most pronounced in the 1^st^ trimester (adjusted OR=1.64, 95% CI:1.45-1.85), though it remained significant in the 2^nd^ trimester as well (adjusted OR=1.48, 95% CI:1.29-1.69). Analysis of specific WBC subtypes revealed a similar association of high levels of neutrophils, eosinophils, and basophiles with increased ASD odds while levels of lymphocytes and monocytes showed a dose-response effect with low and high levels associated with decreased and increased ASD odds respectively **(Table 2)**.

**Table 2:** Association between maternal WBC counts and ASD in the offspring.

| WBC type | WBC levels<br>(Ref- Normal)<sup>a</sup> | Trimester | Crude Odds Ratio<br>(95% CI) | P-value | Adjusted Odds Ratio*<br>(95% CI) | P-value |
|---|---|---|---|---|---|---|
| **Total WBC count** | High<sup>b</sup> | 1<sup>st</sup> or 2<sup>nd</sup> | 1.74 (1.58-1.91) | **<0.001** | 1.59 (1.43-1.75) | **<0.001** |
|  |  | 1<sup>st</sup> | 1.76 (1.57-1.97) | **<0.001** | 1.64 (1.45-1.85) | **<0.001** |
|  |  | 2<sup>nd</sup> | 1.63 (1.43-1.84) | **<0.001** | 1.48 (1.29-1.69) | **<0.001** |
|  | Low<sup>c</sup> | 1<sup>st</sup> or 2<sup>nd</sup> | 0.93 (0.82-1.06) | 0.311 | 0.99 (0.86-1.13) | 0.924 |
|  |  | 1<sup>st</sup> | 0.99 (0.85-1.15) | 0.974 | 1.06 (0.90-1.23) | 0.446 |
|  |  | 2<sup>nd</sup> | 0.82 (0.69-0.97) | **0.031** | 0.86 (0.72-1.03) | 0.124 |
| **Neutrophils** | High<sup>b</sup> | 1<sup>st</sup> or 2<sup>nd</sup> | 1.60 (1.45-1.76) | **<0.001** | 1.48 (1.34-1.64) | **<0.001** |
|  |  | 1<sup>st</sup> | 1.52 (1.34-1.72) | **<0.001** | 1.43 (1.25-1.62) | **<0.001** |
|  |  | 2<sup>nd</sup> | 1.68 (1.48-1.90) | **<0.001** | 1.57 (1.37-1.79) | **<0.001** |
|  | Low<sup>c</sup> | 1<sup>st</sup> or 2<sup>nd</sup> | 0.97 (0.86-1.10) | 0.737 | 1.01 (0.88-1.15) | 0.842 |
|  |  | 1<sup>st</sup> | 1.02 (0.88-1.18) | 0.721 | 1.06 (0.90-1.24) | 0.429 |
|  |  | 2<sup>nd</sup> | 0.93 (0.79-1.08) | 0.379 | 0.97 (0.81-1.14) | 0.73 |
| **Lymphocytes** | High<sup>b</sup> | 1<sup>st</sup> or 2<sup>nd</sup> | 1.49 (1.35-1.65) | **<0.001** | 1.38 (1.23-1.53) | **<0.001** |
|  |  | 1<sup>st</sup> | 1.50 (1.33-1.68) | **<0.001** | 1.42 (1.25-1.61) | **<0.001** |
|  |  | 2<sup>nd</sup> | 1.35 (1.17-1.55) | **<0.001** | 1.24 (1.06-1.43) | **0.003** |
|  | Low<sup>c</sup> | 1<sup>st</sup> or 2<sup>nd</sup> | 0.83 (0.72-0.95) | **0.009** | 0.85 (0.74-0.98) | **0.037** |
|  |  | 1<sup>st</sup> | 0.83 (0.70-0.97) | **0.031** | 0.85 (0.72-1.01) | 0.086 |
|  |  | 2<sup>nd</sup> | 0.85 (0.72-0.99) | 0.054 | 0.89 (0.75-1.05) | 0.196 |
| **Monocytes** | High<sup>b</sup> | 1<sup>st</sup> or 2<sup>nd</sup> | 1.39 (1.25-1.54) | **<0.001** | 1.35 (1.20-1.51) | **<0.001** |
|  |  | 1<sup>st</sup> | 1.28 (1.12-1.46) | **<0.001** | 1.28 (1.11-1.47) | **<0.001** |
|  |  | 2<sup>nd</sup> | 1.38 (1.22-1.55) | **<0.001** | 1.33 (1.16-1.51) | **<0.001** |
|  | Low<sup>c</sup> | 1<sup>st</sup> or 2<sup>nd</sup> | 0.66 (0.53-0.81) | **<0.001** | 0.68 (0.54-0.84) | **<0.001** |
|  |  | 1<sup>st</sup> | 0.83 (0.75-0.91) | **<0.001** | 0.86 (0.77-0.95) | **0.005** |
|  |  | 2<sup>nd</sup> | 0.83 (0.74-0.92) | **0.001** | 0.83 (0.73-0.93) | **0.001** |
| **Eosinophils** | High<sup>b</sup> | 1<sup>st</sup> or 2<sup>nd</sup> | 1.34 (1.17-1.53) | **<0.001** | 1.38 (1.20-1.58) | **<0.001** |
|  |  | 1<sup>st</sup> | 1.22 (1.07-1.38) | **0.001** | 1.22 (1.07-1.40) | **0.002** |
|  |  | 2<sup>nd</sup> | 1.32 (1.15-1.52) | **<0.001** | 1.37 (1.18-1.57) | **<0.001** |
| **Basophils** | Low<sup>c</sup> | 1<sup>st</sup> or 2<sup>nd</sup> | 1.53 (0.995-2.263) | **0.038** | 1.40 (0.88-2.10) | 0.122 |
|  |  | 1<sup>st</sup> | 1.32 (1.21-1.43) | **<0.001** | 1.32 (1.20-1.46) | **<0.001** |
|  |  | 2<sup>nd</sup> | 1.29 (1.15-1.44) | **<0.001** | 1.36 (1.21-1.53) | **<0.001** |

<sup>a</sup> WBC level ≥2.5% or ≤97.5% of the distribution; <sup>b</sup> WBC level <2.5% of the distribution; <sup>c</sup> WBC level >97.5% of the distribution
\*Adjusted for the following socio-economic and clinical variables: (Socio-economic status, child gender, maternal age, maternal BMI, maternal diabetes, maternal hypertension, birth weight, birth week)

### Matched-Sibling Analysis

To account for shared familial confounding, a matched-sibling analysis was performed on a subset of 2,894 sibling pairs with discordant ASD status **(Table 3)**. In this analysis, most of the associations seen in the full cohort were attenuated and did not reach statistical significance except the associations between elevated total WBC counts in the 1^st^ trimester with increased ASD odds (adjusted OR=1.63, 95% CI:1.02-2.59), and low monocytes in the 2^nd^ trimester with decreased ASD odds (adjusted OR=0.68, 95% CI:0.48-0.98). Interestingly, significant within-mother correlations were observed in WBC counts across different pregnancies **(Figure S4 and Figure S5)**, suggesting that maternal WBC levels were relatively stable over time. This finding may help explain the discrepancy between the full-cohort and sibling-matched analyses, as limited within-mother variability reduces the ability of sibling comparisons to detect an association. The findings may also indicate that the association between elevated maternal WBC counts and offspring ASD observed in the full cohort reflects a stable maternal inflammatory or immune condition rather than an acute immune response.

**Table 3:** Results of the matched-siblings analysis.

| WBC type | WBC levels<br>(Ref- Normal)<sup>a</sup> | Trimester | Crude Odds Ratio<br>(95% CI) | P-value | Adjusted Odds<br>Ratio* (95% CI) | P-value |
|---|---|---|---|---|---|---|
| **Total WBC count** | High<sup>b</sup> | 1<sup>st</sup> or 2<sup>nd</sup> | 1.11 (0.83-1.47) | 0.471 | 1.10 (0.78-1.56) | 0.571 |
|  |  | 1<sup>st</sup> | 1.51 (1.02-2.22) | **0.036** | 1.63 (1.02-2.59) | **0.039** |
|  |  | 2<sup>nd</sup> | 0.72 (0.48-1.09) | 0.125 | 0.70 (0.42-1.17) | 0.177 |
|  | Low<sup>c</sup> | 1<sup>st</sup> or 2<sup>nd</sup> | 0.66 (0.44-0.98) | **0.04** | 0.73 (0.46-1.17) | 0.2 |
|  |  | 1<sup>st</sup> | 0.68 (0.42-1.08) | 0.106 | 0.98 (0.57-1.69) | 0.956 |
|  |  | 2<sup>nd</sup> | 0.44 (0.25-0.79) | **0.006** | 0.45 (0.23-0.87) | **0.019** |
| **Neutrophils** | High<sup>b</sup> | 1<sup>st</sup> or 2<sup>nd</sup> | 1.19 (0.90-1.57) | 0.208 | 1.15 (0.82-1.61) | 0.394 |
|  |  | 1<sup>st</sup> | 1.29 (0.90-1.89) | 0.152 | 1.33 (0.88-2.02) | 0.172 |
|  |  | 2<sup>nd</sup> | 1.04 (0.69-1.55) | 0.838 | 1.00 (0.60-1.67) | 0.989 |
|  | Low<sup>c</sup> | 1<sup>st</sup> or 2<sup>nd</sup> | 0.89 (0.61-1.30) | 0.571 | 0.68 (0.43-1.07) | 0.103 |
|  |  | 1<sup>st</sup> | 1.02 (0.64-1.62) | 0.907 | 0.89 (0.51-1.54) | 0.69 |
|  |  | 2<sup>nd</sup> | 0.77 (0.46-1.27) | 0.311 | 0.59 (0.31-1.10) | 0.101 |
| **Lymphocytes** | High<sup>b</sup> | 1<sup>st</sup> or 2<sup>nd</sup> | 1.04 (0.78-1.39) | 0.768 | 0.92 (0.64-1.31) | 0.66 |
|  |  | 1<sup>st</sup> | 1.42 (1.01-2.01) | **0.041** | 1.35 (0.90-2.02) | 0.138 |
|  |  | 2<sup>nd</sup> | 0.80 (0.53-1.21) | 0.303 | 0.65 (0.39-1.07) | 0.096 |
|  | Low<sup>c</sup> | 1<sup>st</sup> or 2<sup>nd</sup> | 0.72 (0.52-1.01) | 0.057 | 0.80 (0.53-1.19) | 0.276 |
|  |  | 1<sup>st</sup> | 0.77 (0.51-1.16) | 0.222 | 0.88 (0.53-1.46) | 0.637 |
|  |  | 2<sup>nd</sup> | 0.65 (0.44-0.96) | **0.03** | 0.81 (0.51-1.28) | 0.381 |
| **Monocytes** | High<sup>b</sup> | 1<sup>st</sup> or 2<sup>nd</sup> | 1.30 (0.96-1.76) | 0.081 | 1.11 (0.76-1.62) | 0.566 |
|  |  | 1<sup>st</sup> | 1.40 (0.96-2.04) | 0.075 | 1.25 (0.79-1.98) | 0.324 |
|  |  | 2<sup>nd</sup> | 0.90 (0.61-1.33) | 0.619 | 0.80 (0.49-1.28) | 0.36 |
|  | Low<sup>c</sup> | 1<sup>st</sup> or 2<sup>nd</sup> | 0.67 (0.42-1.08) | 0.101 | 0.89 (0.50-1.56) | 0.69 |
|  |  | 1<sup>st</sup> | 0.75 (0.58-0.96) | **0.027** | 0.79 (0.58-1.06) | 0.121 |
|  |  | 2<sup>nd</sup> | 0.69 (0.51-0.93) | **0.015** | 0.68 (0.48-0.98) | **0.042** |
| **Eosinophils** | High<sup>b</sup> | 1<sup>st</sup> or 2<sup>nd</sup> | 1.09 (0.71-1.67) | 0.666 | 1.17 (0.72-1.90) | 0.514 |
|  |  | 1<sup>st</sup> | 0.91 (0.60-1.37) | 0.673 | 0.91 (0.56-1.47) | 0.714 |
|  |  | 2<sup>nd</sup> | 1.28 (0.80-2.06) | 0.288 | 1.8 (1.02-3.18) | **0.040** |
| **Basophils** | Low<sup>c</sup> | 1<sup>st</sup> or 2<sup>nd</sup> | 1 (0.28-3.45) | 1 | 0.52 (0.10-2.61) | 0.43 |
|  |  | 1<sup>st</sup> | 0.76 (0.54-1.068) | 0.115 | 0.63 (0.42-0.94) | **0.024** |
|  |  | 2<sup>nd</sup> | 1.04 (0.70-1.54) | 0.822 | 0.96 (0.61-1.51) | 0.865 |

<sup>a</sup> WBC level ≥2.5% or ≤97.5% of the distribution; <sup>b</sup> WBC level <2.5% of the distribution; <sup>c</sup> WBC level >97.5% of the distribution
\*Adjusted for the following socio-economic and clinical variables: (Socio-economic status, child gender, maternal age, maternal BMI, maternal diabetes, maternal hypertension, birth weight, birth week)

### Chronicity Analyses

To examine whether the maternal WBC levels associated with offspring’s ASD reflect an acute or chronic inflammatory condition, we used the sub-cohort of multiparous mothers in the study by comparing the odds of ASD among mothers with consistent inter-pregnancy elevation in their WBC counts to their counterparts. The results of this chronicity analysis are summarized in **Table 4**. Persistent elevation in maternal total WBC counts was significantly associated with increased odds of ASD in offspring using either the 97.5^th^- or 95^th^- percentile cutoff (adjusted OR = 1.49, 95% CI: 1.08-2.01, and adjusted OR = 1.5, 95% CI: 1.22-1.82, respectively). Similar associations were observed for most WBC subtypes, except monocytes, further supporting the presence of a broad and stable maternal immune phenotype rather than an isolated elevation in a single leukocyte subtype.

**Table 4:** Results of the chronicity analysis.

| WBC type | Chronicity<br>Threshold<br>(Ref- Normal) | Trimester | Crude Odds Ratio<br>(95% CI) | P-value | Adjusted Odds<br>Ratio* (95% CI) | P-value |
|---|---|---|---|---|---|---|
| **Total WBC<br>count** | Top 2.5% | 1<sup>st</sup> or 2<sup>nd</sup> | 1.70 (1.25-2.26) | **<0.001** | 1.49 (1.08-2.01) | **0.012** |
|  |  | 1<sup>st</sup> | 1.65 (1.07-2.43) | **0.016** | 1.44 (0.91-2.15) | 0.098 |
|  |  | 2<sup>nd</sup> | 1.78 (1.10-2.72) | **0.011** | 1.56 (0.94-2.43) | 0.066 |
|  | Top 5% | 1<sup>st</sup> or 2<sup>nd</sup> | 1.71 (1.40-2.07) | **<0.001** | 1.51 (1.22-1.82) | **<0.001** |
|  |  | 1<sup>st</sup> | 1.71 (1.29-2.21) | **<0.001** | 1.46 (1.08-1.92) | **0.010** |
|  |  | 2<sup>nd</sup> | 1.75 (1.30-2.30) | **<0.001** | 1.54 (1.12-2.05) | **0.004** |
| **Neutrophils** | Top 2.5% | 1<sup>st</sup> or 2<sup>nd</sup> | 1.51 (1.05-2.10) | **0.018** | 1.42 (0.98-1.98) | 0.051 |
|  |  | 1<sup>st</sup> | 1.27 (0.74-2.04) | 0.349 | 1.17 (0.66-1.91) | 0.557 |
|  |  | 2<sup>nd</sup> | 1.92 (1.14-3.03) | **0.008** | 1.85 (1.10-2.93) | **0.013** |
|  | Top 5% | 1<sup>st</sup> or 2<sup>nd</sup> | 1.55 (1.24-1.90) | **<0.001** | 1.39 (1.11-1.73) | **0.003** |
|  |  | 1<sup>st</sup> | 1.70 (1.27-2.23) | **<0.001** | 1.56 (1.15-2.07) | **0.003** |
|  |  | 2<sup>nd</sup> | 1.77 (1.30-2.36) | **<0.001** | 1.53 (1.10-2.07) | **0.008** |
| **Lymphocytes** | Top 2.5% | 1<sup>st</sup> or 2<sup>nd</sup> | 1.82 (1.37-2.37) | **<0.001** | 1.73 (1.29-2.26) | **<0.001** |
|  |  | 1<sup>st</sup> | 1.81 (1.19-2.64) | **0.003** | 1.70 (1.11-2.51) | **0.010** |
|  |  | 2<sup>nd</sup> | 1.55 (0.99-2.32) | **0.046** | 1.50 (0.94-2.28) | 0.068 |
|  | Top 5% | 1<sup>st</sup> or 2<sup>nd</sup> | 1.72 (1.41-2.07) | **<0.001** | 1.64 (1.33-1.99) | **<0.001** |
|  |  | 1<sup>st</sup> | 1.73 (1.33-2.22) | **<0.001** | 1.64 (1.24-2.11) | **<0.001** |
|  |  | 2<sup>nd</sup> | 1.54 (1.12-2.06) | **<0.001** | 1.35 (1.09-2.06) | **0.009** |
| **Monocytes** | Top 2.5% | 1<sup>st</sup> or 2<sup>nd</sup> | 1.53 (1.06-2.14) | **0.169** | 1.33 (0.88-1.93) | 0.154 |
|  |  | 1<sup>st</sup> | 1.06 (0.58-1.78) | 0.836 | 0.81 (0.38-1.48) | 0.526 |
|  |  | 2<sup>nd</sup> | 2.11 (1.21-3.42) | **0.004** | 2.03 (1.12-3.41) | **0.012** |
|  | Top 5% | 1<sup>st</sup> or 2<sup>nd</sup> | 1.21 (0.92-1.55) | 0.160 | 1.06 (0.78-1.40) | 0.709 |
|  |  | 1<sup>st</sup> | 1.18 (0.77-1.73) | 0.418 | 0.96 (0.58-1.50) | 0.867 |
|  |  | 2<sup>nd</sup> | 1.34 (0.92-1.88) | 0.106 | 1.34 (0.91-1.92) | 0.120 |
| **Eosinophils** | Top 2.5% | 1<sup>st</sup> or 2<sup>nd</sup> | 1.62 (1.13-2.26) | **0.006** | 1.72 (1.19-2.41) | **0.002** |
|  |  | 1<sup>st</sup> | 1.36 (0.77-2.22) | 0.245 | 1.36 (0.75-2.26) | 0.266 |
|  |  | 2<sup>nd</sup> | 1.88 (1.15-2.89) | **0.007** | 1.92 (1.16-3.00) | **0.007** |
|  | Top 5% | 1<sup>st</sup> or 2<sup>nd</sup> | 1.44 (1.12-1.81) | **0.003** | 1.44 (1.11-1.84) | **0.004** |
|  |  | 1<sup>st</sup> | 1.44 (1.03-1.97) | **0.026** | 1.41 (0.98-1.95) | **0.048** |
|  |  | 2<sup>nd</sup> | 1.19 (0.80-1.69) | 0.357 | 1.19 (0.79-1.71) | 0.377 |

Chronic exposure was defined as a maternal phenotype exceeding the respective percentile threshold (top 2.5% or top 5.0%) across all recorded pregnancies. The reference group comprised of mothers whose white blood cell (WBC) counts were within the normal range (i.e. WBC level ≥2.5% or ≤97.5% of the distribution, and WBC level ≥5.0% or ≤95.0% respectively) across all tracked pregnancies, requiring a minimum of two valid gestations with available data. Models for basophil counts could not be statistically evaluated due to an insufficient number of mothers meeting the strict chronic exposure criteria, resulting in a lack of model convergence.
\* Adjusted for maternal background characteristics calculated at the maternal level: mean maternal age across all pregnancies, maternal BMI at the first recorded pregnancy, socioeconomic status at the first recorded pregnancy, maternal diabetes mellitus (defined as a diagnosis in at least one pregnancy), and maternal hypertension (defined as a diagnosis in at least one pregnancy).

## Discussion

In this large population-based cohort study comprising over 300,000 pregnancies, we identified a robust association between a persistent (chronic) elevated maternal WBC counts during pregnancy and the risk of ASD in offspring. The association was evident across total WBC counts as well as specific subtypes. Notably, the risk was most pronounced during the 1^st^ trimester, a critical window for fetal neurodevelopment, consistent with other studies highlighting the vulnerability of this period.(2-4) Furthermore, the pattern of associations was directionally consistent with higher risk at elevated values and lower risk at some lower lymphocyte and monocyte values.

Our results align with the well-established Maternal Immune Activation (MIA) hypothesis and its contribution to altered fetal neurodevelopment. (10) While previous studies have focused primarily on maternal infections or specific cytokine levels (14, 16), our results suggest that routinely measured hematological markers may also provide accessible indicators of maternal immune activity associated with ASD risk. The association with both neutrophils (markers of innate immunity) and lymphocytes (adaptive immunity) suggests a generalized state of immune activation rather than a reaction to a specific pathogen. This supports the notion that the intensity of the maternal immune response plays a key role in the pathogenesis of neurodevelopmental deviations. (11)

Collectively, our findings suggest that the association between elevated maternal WBC counts during pregnancy and ASD in offspring may reflect a persistent maternal inflammatory condition rather than an acute immune response. The attenuation of the association in the sibling-matched analysis may be explained by the substantial stability of maternal WBC counts across pregnancies observed in the interpregnancy correlation analyses. Because sibling fixed-effects models rely exclusively on within-mother variation in exposure, limited variation across pregnancies may reduce their ability to detect associations, particularly when mothers with consistently elevated inflammatory profiles expose multiple offspring to similar prenatal immune environments. This interpretation is further supported by the chronicity analysis, which showed an increased risk of ASD among offspring of mothers with persistently elevated WBC counts.

### Strengths and Limitations

The study’s main strength is its large sample size and the use of unselected, real-world data from a unified healthcare provider. The included cohort represents a broad cross-section of the Jewish population in Israel, encompassing individuals from diverse geographic regions and socioeconomic backgrounds, supporting the generalizability of the findings within this population. Additionally, the availability of rich electronic health records allowed for extensive adjustment for a wide range of maternal and perinatal covariates.

While our study offers important insights, several limitations should be acknowledged. First, certain unmeasured confounders were unavailable or incompletely captured in our dataset. These include paternal age, maternal smoking, acute fever, gestational antibiotic use, and specific medications such as corticosteroids or immunomodulators. Furthermore, granular data on maternal psychiatric history, underlying autoimmune diseases, or overall healthcare utilization intensity were lacking. The absence of these variables may contribute to residual confounding of our findings. For example, unmeasured chronic maternal conditions or maternal smoking could independently drive both elevated leukocyte counts and altered fetal neurodevelopment, potentially inflating estimates in our population-level models. Crucially, our matched-sibling analysis explicitly mitigates these concerns. By contrasting discordant siblings, the fixed-effects design naturally controls for all stable, unmeasured genetic factors, maternal lifestyle traits, and underlying healthcare-seeking behaviors that remain constant across pregnancies. Another limitation relates to exposure timing. Because maternal WBC counts were obtained as part of routine clinical care, the exact gestational age at the time of testing varied among participants. Given the dynamic nature of immune function throughout gestation, this variability may introduce a degree of temporal misclassification. However, such random variation typically introduces noise that attenuates rather than inflates observed associations. Consequently, this limitation most likely biases our estimates toward the null, suggesting that our findings represent a conservative underestimate of the true biological relationship. Finally, WBC counts are non-specific inflammatory markers. Elevated WBC levels can reflect diverse underlying conditions, from subclinical metabolic processes to acute infections. While our findings establish a robust epidemiological link, future research incorporating specific immune biomarkers (e.g., cytokines, CRP) is warranted to dissect the precise underlying biological pathways. Therefore, causal inferences of our findings should be made cautiously.

## Conclusions and Future Directions

The findings of this study indicate that persistently elevated maternal WBC counts, which may reflect chronic immune dysregulation even at non-pathological levels, can influence fetal neurodevelopment through immune-mediated mechanisms. While immediate translation into standalone clinical screening is premature, our study provides further support to the maternal immune activation hypothesis within a massive, real-world population. Ultimately, pending external validation and mechanistic clarification, these readily accessible hematologic trajectories could hold significant public health value, potentially contributing to future multifactorial prenatal risk-assessment frameworks

## Author Contributions

AT and AD contributed equally to this work. AT, AD, IM and GM conceived and designed the study. AT, and AD performed the statistical analysis and interpreted the data. GM and IM provided the resources and data for the study. AT drafted the manuscript. IM critically revised the manuscript for important intellectual content. All authors gave final approval of the version to be submitted and agree to be accountable for all aspects of the work.

## Data Availability

The data that support the findings of this study are available from Clalit Health Services (CHS), but restrictions apply to the availability of these data, which were used under license for the current study, and so are not publicly available due to patient confidentiality and institutional data regulations.

