## Supplementary Tables and Figures for "Chronic Maternal White Blood Cell Elevation During Pregnancy and Offspring Autism"

**Supplementary Information:**
Table S1: Absolute numeric cut-points for maternal WBC counts by gestational trimester:

| Cell Type | Gestational Trimester | Low Cut-point  (≤2.5th percentile)  In cells/µL X 10^3^ | High Cut-point  (≥97.5th percentile)  In cells/µL X 10^3^ |
| --- | --- | --- | --- |
| Total WBC count | 1^st^ | 4.01 | 11.60 |
|  | 2^nd^ | 4.70 | 13.10 |
| Neutrophils | 1^st^ | 2.01 | 8.20 |
|  | 2^nd^ | 2.90 | 10.00 |
| Lymphocytes | 1^st^ | 1.01 | 3.30 |
|  | 2^nd^ | 0.90 | 2.93 |
| Monocytes | 1^st^ | 0.20 | 0.76 |
|  | 2^nd^ | 0.20 | 0.80 |
| Eosinophils* | 1^st^ | 0.00 | 0.50 |
|  | 2^nd^ | 0.00 | 0.50 |
| Basophils* | 1^st^ | 0.00 | 0.10 |
|  | 2^nd^ | 0.00 | 0.10 |

*** For eosinophils and basophils, the 2.5th percentile lower boundary equals 0.00. This floor effect is a recognized physiological and technological characteristic in large-scale epidemiological cohorts due to the low baseline circulating frequency of these specific leukocyte subpopulations in healthy individuals, resulting in a true absolute zero count within the bottom quantiles of the distribution.

Table S2: Sociodemographic, maternal, and delivery characteristics of the outcome groups- autism diagnosis in offspring:

| Variable | | | ASD offspring  (N=8,106) | | Non- ASD offspring  (N= 295,128) | P-value | | |
| --- | --- | --- | --- | --- | --- | --- | --- | --- |
| Socio-demographic characteristics | | | | | | | | |
| Maternal age [mean (+ SD)] | | | 32.3 (±5.3) | | 31.6 (±5.1) | | | <0.001 |
| Offspring sex  [N(%)] | **Male** | 6,150 (76%) | | 149,562 (51%) | | | <0.001 | |
|  | **Female** | 1,956 (24%) | | 145,566 (49%) | | |  |  |
| Socioeconomic status  [N(%)] | **Very low** | 27 (0.4%) | | 3,578 (1.3%) | | | <0.001 | |
|  | **Low** | 1,513 (20%) | | 57,559 (21%) | | |  |  |
|  | **Medium** | 3,081 (41%) | | 103,555 (37%) | | |  |  |
|  | **High** | 2,377 (31%) | | 87,766 (32%) | | |  |  |
|  | **Very high** | 607 (8.0%) | | 25,278 (9.1%) | | |  |  |
| Maternal characteristics | | | | | | | | |
| Maternal BMI [mean (+ SD)] | | | 22.9 (±5.0) | | 22.1 (±4.3) | | | <0.001 |
| Gestational diabetes:  [N(%)] | | | 746 (9.2%) | | 15,549 (5.3%) | | | <0.001 |
| Gestational hypertension:  [N(%)] | | | 504 (6.2%) | | 10,388 (3.5%) | | | <0.001 |
| Gestational anemia:  [N(%)] | | | 1,354 (17%) | | 44,706 (15%) | | | <0.001 |
| IVF:  [N(%)] | | | 428 (5.3%) | | 11,105 (3.8%) | | | <0.001 |
| Delivery characteristics | | | | | | | | |
| Birth weight (gr) [mean (+ SD)] | | | 3,252 (±473) | | 3,284 (±434) | | | 0.001 |
| Birth week [mean (+ SD)] | | | 38.88 (±1.54) | | 39.15 (±1.41) | | | <0.001 |

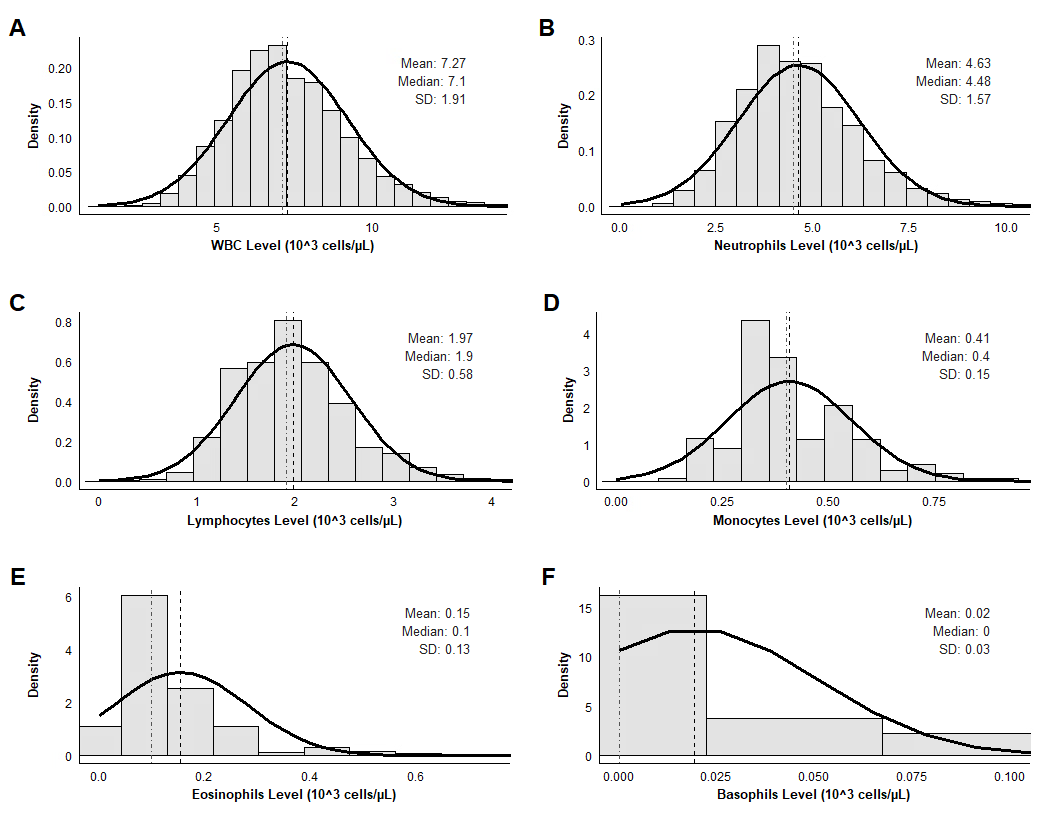
Figure S1: Histograms of maternal WBC and subtype distributions during the 1^st^ trimester:

Figure S1: Histograms of maternal WBC and subtype distributions during the 1^st^ trimester- Population-level density distributions and frequency histories obtained during the 1^st^ trimester of pregnancy (weeks 0 to 13+6) for: **(A)** total white blood cell (WBC) counts, **(B)** neutrophils, **(C)** lymphocytes, **(D)** monocytes, **(E)** eosinophils, and **(F)** basophils. The solid black curve represents the overlaid theoretical normal distribution line. The vertical dashed line indicates the empirical cohort mean, and the vertical dot-dashed line indicates the empirical cohort median. Computed standard deviations (SD) and summary statistics are displayed in the upper-right corner of each panel. Units are expressed in 10^3 cells/µL.

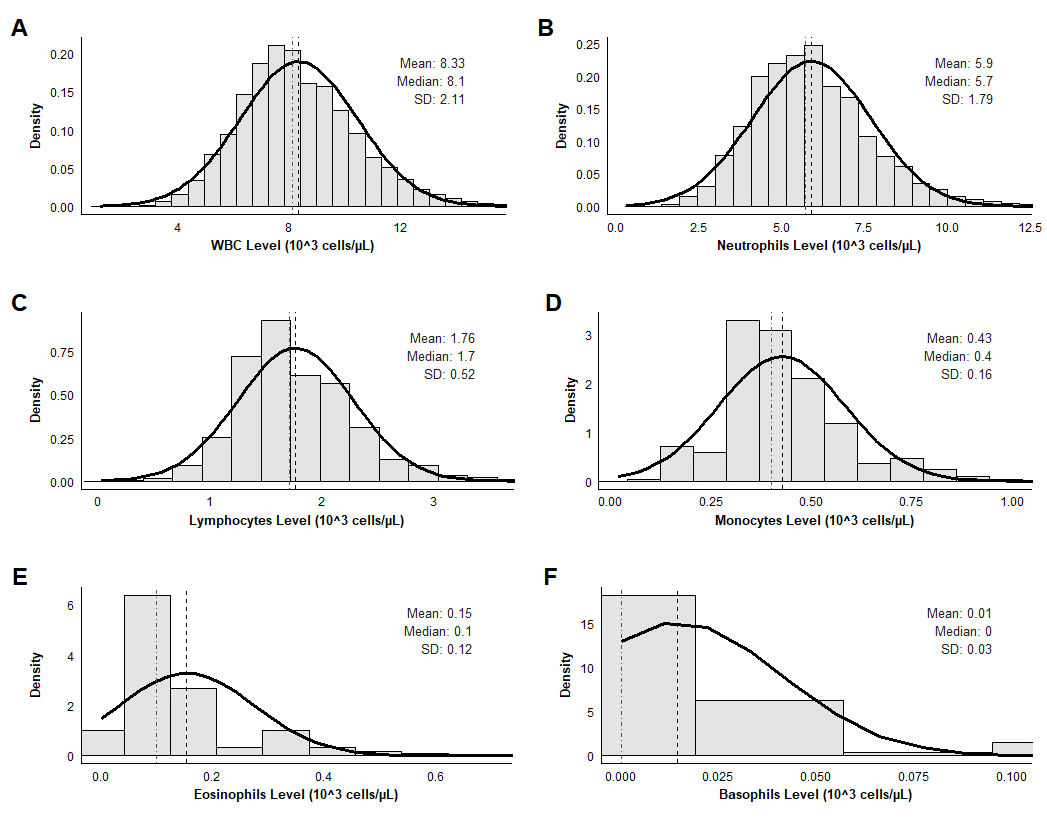
Figure S2: Histograms of maternal WBC and subtype distributions during the 2^nd^ trimester

Figure S2: Histograms of maternal WBC and subtype distributions during the 2^nd^ trimester- Population-level density distributions and frequency histories of maternal total white blood cell (WBC) counts, lymphocytes, neutrophils, monocytes, basophils, and eosinophils obtained during the 2^nd^ trimester of pregnancy (weeks 14 to 26+6). The solid black curve represents the overlaid theoretical normal distribution line. The vertical dashed line indicates the empirical cohort mean, and the vertical dot-dashed line indicates the empirical cohort median. Computed standard deviations (SD) and summary statistics are displayed in the upper-right corner of each panel. Units are expressed in 10^3 cells/µL.

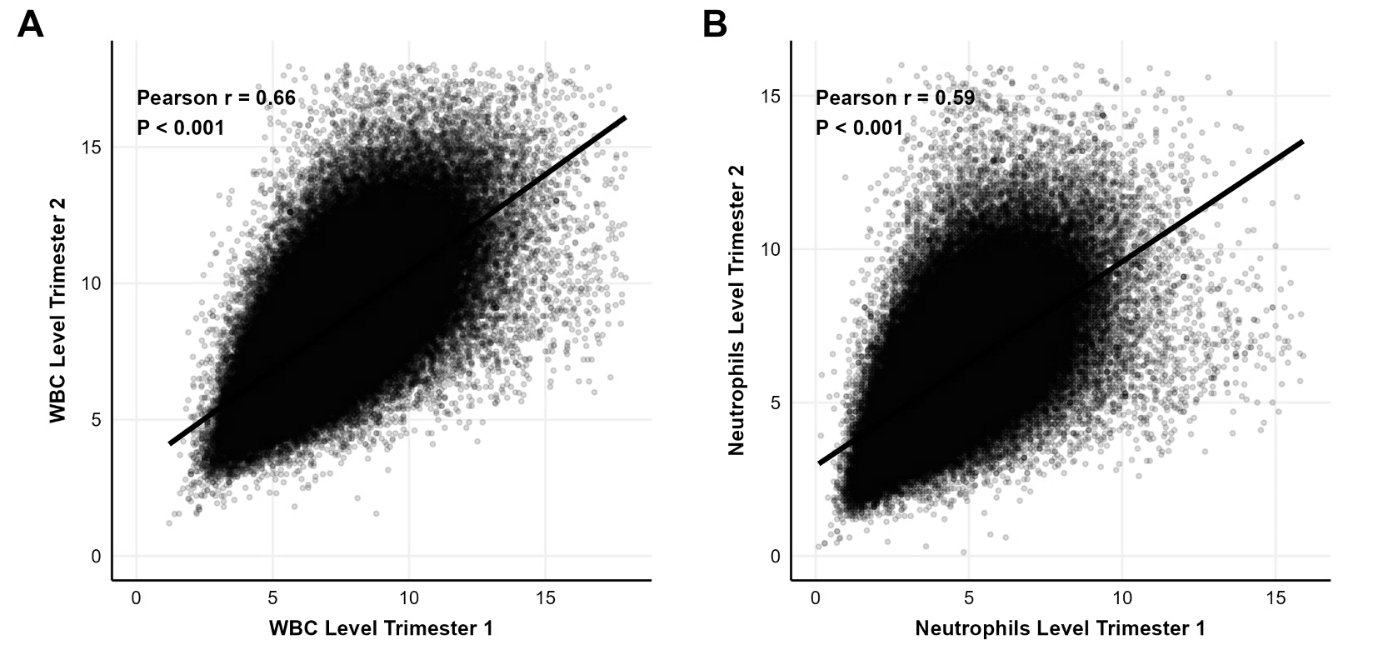
Figure S3: Scatterplots showing correlations of WBC counts between the 1^st^ and 2^nd^ trimesters:

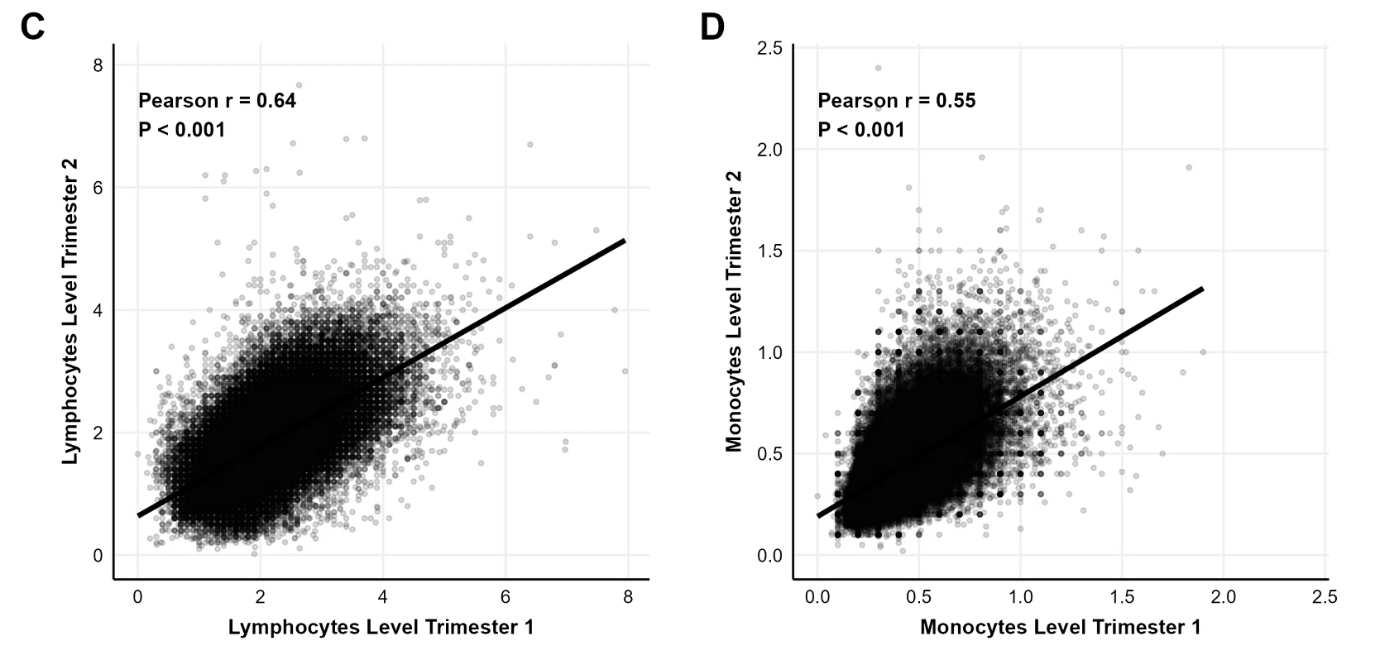

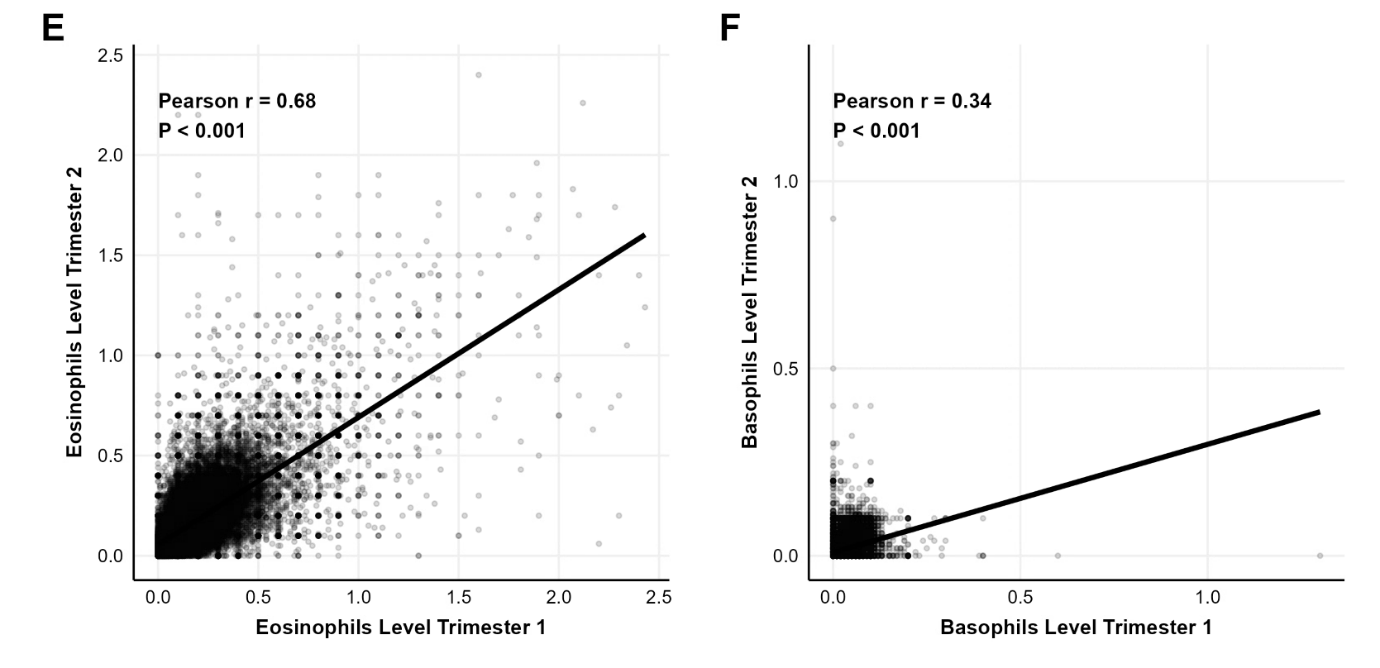

Figure S3: Scatterplots showing correlations of WBC counts between the 1^st^ and 2^nd^ trimesters- Longitudinal tracking and intra-individual exposure stability of maternal absolute leukocyte profiles across different phases of gestation. Scatterplots display the direct linear correlation between 1^st^ (x-axis) and 2^nd^ (y-axis) trimesters laboratory metrics for: **(A)** total WBC counts, **(B)** neutrophils, **(C)** lymphocytes, **(D)** monocytes, **(E)** eosinophils, and **(F)** basophils. Solid black lines represent fitted linear regression slopes, evaluated using Pearson correlation coefficients (r) and corresponding two-tailed P-values displayed in each panel. Units are expressed in 10^3 cells/µL.

Figure S4: Cross-sibling correlation of maternal 1^st^ trimester WBC/subtypes
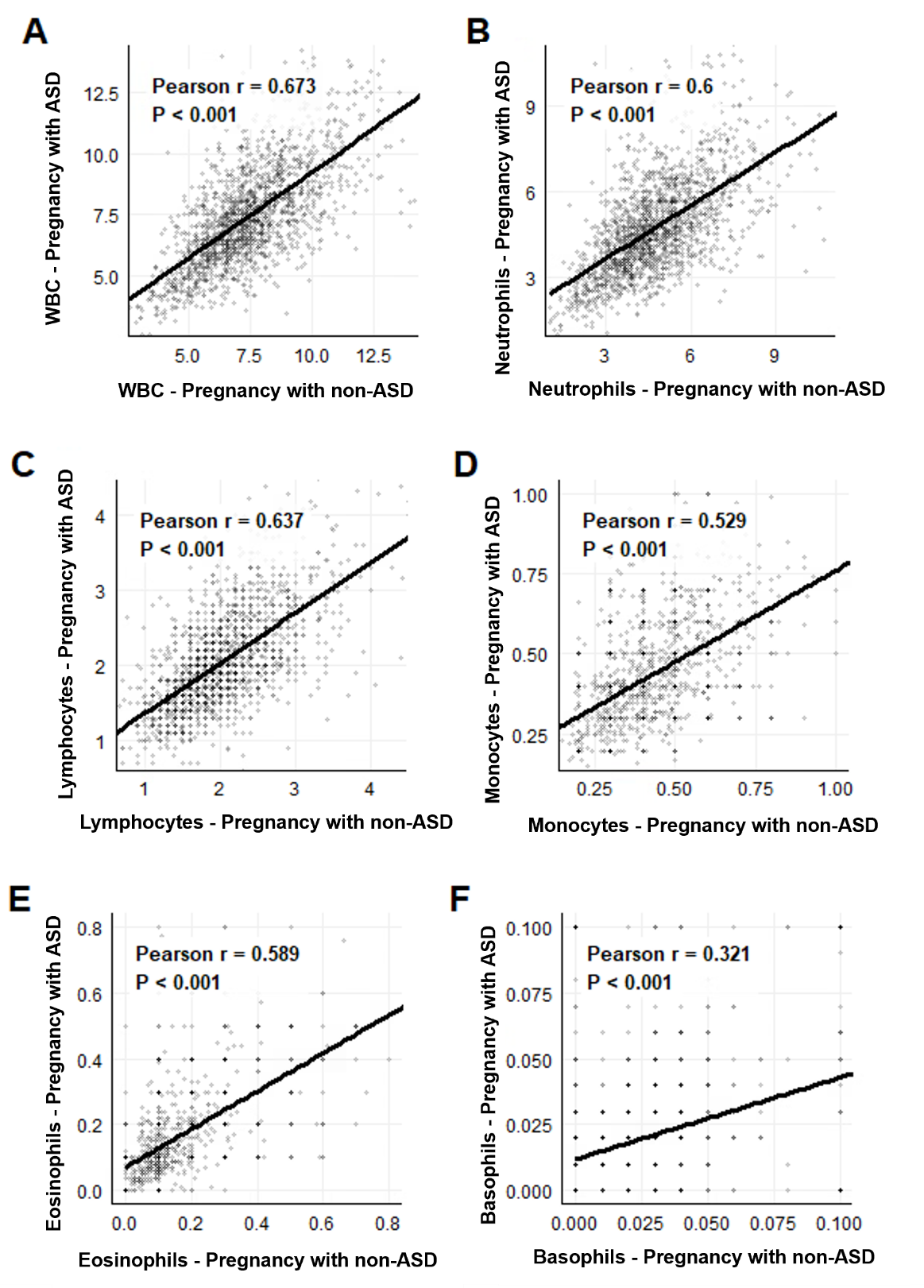
(ASD vs non-ASD):

Figure S4: Cross-sibling correlation of maternal 1^st^ trimester WBC and cell subtypes- Familial exposure clustering and trait stability between separate pregnancies within the sibling sub-cohort during early gestation. Bivariate scatterplots display the correlation of maternal 1^st^ trimester absolute counts when carrying a child subsequently diagnosed with ASD (y-axis) compared to an unaffected sibling (x-axis) for: **(A)** total WBC, **(B)** neutrophils, **(C)** lymphocytes, **(D)** monocytes, **(E)** eosinophils, and **(F)** basophils. Solid black lines denote linear regression fits, evaluated via Pearson correlation coefficients (r) and two-tailed p-values. Units are expressed in 10^3 cells/µL.

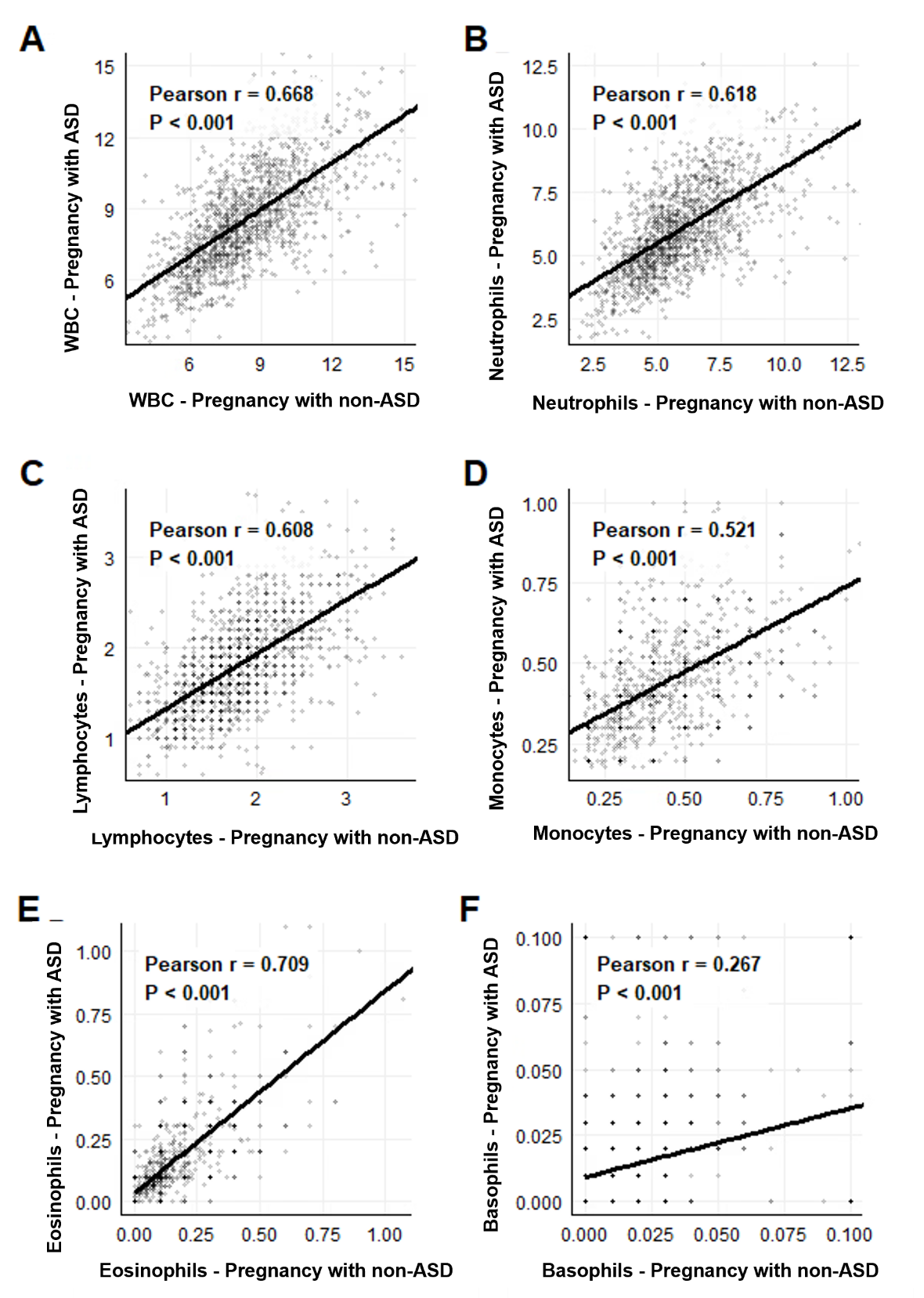
Figure S5: Cross-sibling correlation of maternal 2^nd^ trimester WBC/subtypes (ASD vs non-ASD):

Figure S5: Cross-sibling correlation of maternal 2^nd^ trimester WBC and cell subtypes- Familial exposure clustering and trait stability between separate pregnancies within the sibling sub-cohort during later gestation. Bivariate scatterplots display the correlation of maternal 2^nd^ trimester absolute counts when carrying a child subsequently diagnosed with ASD (y-axis) compared to an unaffected sibling (x-axis) for: **(A)** total WBC, **(B)** neutrophils, **(C)** lymphocytes, **(D)** monocytes, **(E)** eosinophils, and **(F)** basophils. Solid black lines denote linear regression fits, evaluated via Pearson correlation coefficients (r) and two-tailed p-values. Units are expressed in 10^3 cells/µL.
